# A Rapid Risk Assessment for Measles Outbreaks in North Carolina

**DOI:** 10.1101/2025.09.10.25335511

**Authors:** Michael E. DeWitt, Brinkley Raynor Bellotti, Nicholas Kortessis, Avinash K. Shetty, Julianne Green, Anupama Neelakanta, Areej Bukhari, Amina Ahmed, Werner Bischoff, Catherine Passaretti

## Abstract

**Background:** On June 24, 2025, the North Carolina Department of Health and Human Services (NCDHHS) announced a measles case from an international traveler which resulted in multiple exposures including attendance at the Greensboro Science Center, which hosts over 621,000 visitors annually. Given declining measles vaccination rates across the US and ongoing national outbreaks, we estimated potential secondary cases that might occur if measles were introduced into different elementary schools in North Carolina.

**Methods:** We used kindergarten immunization data from NCDHHS (2019–2023) to estimate potential secondary cases using a Susceptible, Exposed, Infected, Recovered, Vaccinated (SEIRV) model. We then estimated the marginal impact of vaccination on outbreak risk.

**Results:** Only 47 of North Carolina’s 100 counties have the recommended full vaccination rates of at least 95%. We found that over 41,000 out of over 641,000 (6.5%) elementary-aged students have not been fully vaccinated against the measles. Of the 1,474 elementary schools we considered, 142 elementary schools have a greater than 50% probability of at least 10 cases should measles be introduced; 28 schools have probabilities exceeding 80%.

**Limitations:** We assume that the SEIRV framework adequately captures the dynamics of students within a school with homogeneous mixing.

**Conclusions:** Measles vaccination coverage in North Carolina has decreased, leaving many children at risk for infection. Many schools have vaccination levels low enough to cause large, sustained outbreaks if a single measles case were introduced. By providing these approximate, order-of-magnitude estimates to public health officials, they can adequately plan for outbreak response and prioritize school and communities at risk for vaccination campaigns.

## Introduction

Several large outbreaks of measles in the United States have been reported in 2025, initially among communities in New Mexico, Oklahoma, and Texas with low vaccination rates[1]. As of September 2, 1413 cases of measles have been reported in 2025 by 42 jurisdictions in the United States—and the worst year since measles was declared eradicated in 2000[2] and coinciding with declining trends in measles-mumps-rubella (MMR) vaccination across the United States[3]. On June 24, 2025, the North Carolina Department of Health and Human Services (NCDHHS) announced the detection of a case of measles from an international traveler[4]. This individual visited several high exposure areas, including an international airport, a children’s science center, and multiple restaurants and grocery stores, over the course of two days and spanning two counties. The individual’s visit to the Greensboro Science Center was a major concern given the high-risk location for measles exposure. It reported over 621,000 visitors in 2023-2024, an average of 1,702 visitors per day over a single year[5]—an astounding number of potential exposures given that the measles virus can linger in the air for up to 2 hours[6] and remain on surfaces for 5-8 days[7].

North Carolina, like many other locations across the United States[3,8], has seen the number of reported kindergarteners with the measles vaccines decline in recent years, from 94.2% in 2020 to 92.5% in 2024 [9]. Due to the highly transmissible nature of the measles virus, with estimates of the basic reproduction number, R_0_, between 12 and 18[10], there is substantial risk of outbreaks even with marginal reductions in population level vaccination rates. Research points out that population level measles vaccination rates must remain above 95% to avoid an outbreak[11]. With falling vaccination rates, the likelihood of onward transmission from a single infected individual rapidly increases. Furthermore, 30% of those individuals who are unvaccinated and infected with the measles virus go on to have complications from disease[12]. While postexposure prophylaxis with vaccine or immunoglobulins can be effective for high-risk individuals, the narrow time windows for administration (72 hours for vaccine, 6 days for immunoglobulin)[12], in addition to limited availability, create significant operational challenges for public health officials and health systems that limit their practical implementation.

To guide the public health response and to understand the risk for outbreaks within different contexts, it is essential to quantify the at-risk population. School aged children represent the primary source of measles transmission in the United States[13]. In this analysis we estimated the number of under and unvaccinated elementary aged students at risk for measles in North Carolina at both the county level and elementary school levels using data made available by NCDHHS. This information can be used by public health departments and the general population to quantify the potential risk of outbreaks in different settings, prioritize schools and communities for vaccination campaigns, and pre-position healthcare materials and resources for rapid deployment when cases occur.

## Methods

### Data Sources

The measles vaccine series is a two-dose series with the first dose occurring generally after one year of age and the second dose occurring a minimum of 28 days later but generally administered between ages 4 and 6 [14]. Fully vaccinated is defined as documentation of a two doses of the MMR vaccine series while partial vaccination is defined as a single dose in the series[14].To estimate the population and vaccination status of elementary aged children by county in 2025, we extracted data on the number of children categorized by NCDHHS as unvaccinated, partially vaccinated, fully vaccinated or provisionally enrolled (i.e., on a doctor certified immunization catch-up plan or receiving vaccine doses at the minimum medically approved intervals) at both the county level and by elementary school within each county for North Carolina using data from the NCDHHS Kindergarten Immunization Dashboard[9]. These data were available for the 2020-2021, 2021-2022, and 2023-2024 school years. School names were then harmonized due to year-to-year differences in spelling (e.g., addition of “ School” and abbreviations). Elementary schools are typically composed of kindergarten through the 5^th^ grade meaning that this data dashboard does not include vaccination information for children in kindergarten and 5^th^ grade during the 2024-2025 school year. To fill these gaps, we assumed the 2020-2021 data represents the 2019-2020 values (representing current 5^th^ grade students) and the 2023-2024 values represent 2024-2025 values (representing the current kindergarten students). We assumed that at the time of writing those students provisionally enrolled in vaccination catch-up programs for kindergarten were still susceptible to infection. Furthermore, when analyzing individual elementary school level data, we included those schools with at least 3 or more years of data.

Counties were classified using the North Carolina Rural Center’s definition which considers the 2020 Decennial Census population estimates and defines rural counties as those with 250 people or fewer per square mile [15].

### Outbreak risk model

We created a stochastic compartmental model to represent the transmission dynamics of measles within different contexts and population sizes. Briefly, we divided the elementary school population into Susceptible (those who may be infected with the measles virus), Exposed, Infected, Recovered, and Vaccinated individuals (Figure 1). The stochastic analogue of the model shown in Figure 1 allows us to calculate the number of children transitioning between each state using draws from binomial distributions. Rate parameters are converted to probabilities of transitioning between states (e.g., the probability of a child moving from exposed to infected occurs at 1 – exp(-σ dt), where dt represents the size of the time step). We assumed parameter values as shown in Table 1 and then modeled the total number of new infections in a given population assuming the introduction of a single, unvaccinated, infected individual. While infectious duration may be as long as 8-10 days[12,16], we assume that any child with the presence of rash and fever by day 5 will be moved to isolation and no longer within the closed, school environment thus representing the effective “ recovery” rate. A portion of vaccinated individuals were considered susceptible as per the vaccine efficiencies determined for each dose. However, infectious duration was substantially shortened for those who were considered vaccinated and infected[14]. Additionally, we assumed that children who were partially vaccinated received the one dose after their first birthday. Note that the contact rate was calculated as R_0_ multiplied by the recovery rate. We ran the stochastic model run 1000 times using the odin package[17] for each county and elementary school where there were at least 3 years of vaccination data available. From these runs, we calculated the average and 95% quantiles for the number of new infections along with the proportion of scenarios in which 10 or more individuals were infected, taking this value to represent the probability of an outbreak.

**Table 1.** Parameter values used in the stochastic SEIRV model.

| Parameter | Value |
| --- | --- |
| Reproduction Number, $R_0$ | 15 [10] |
| Contact rate, $\beta$ | 3.0 (calculated as $R_0 * \gamma$) |
| Incubation rate, $\sigma$ | 1/10 days[18] |
| Recovery rate (infectiousness) before isolation, $\gamma$ | 1/5 days [12,16] |
| Vaccine effectiveness (1 Dose) | 93%[14] |
| Vaccine effectiveness (2 Doses) | 97%[14] |
| Reduction in infectiousness among infected and vaccinated | 95%[19–21] |

**Figure 1.**
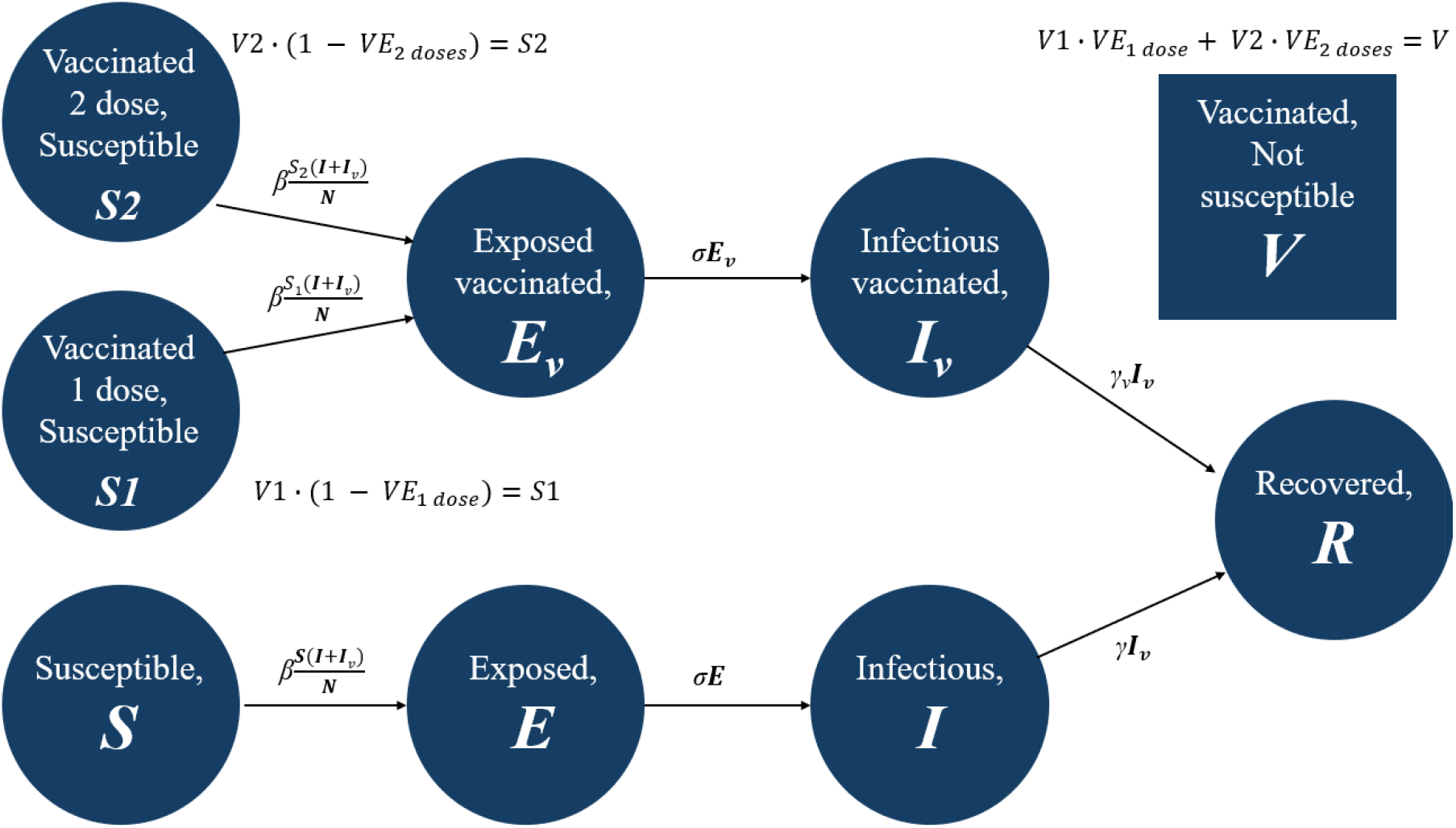
Susceptible, Exposed, Infected, Recovered, Vaccinated compartment model for the spread of measles within a closed population. Note that demographic effects are not included (i.e., births and deaths).

Given the fast dynamics of the measles outbreaks (e.g., high infectiousness) we did not include any demographic effects (i.e., births, deaths), nor did we model waning antibody titers among the vaccinated or recovered, at least over the duration of the outbreaks.

### Exposure modeling

To estimate the probability that an under vaccinated or unvaccinated person encounters an infected individual in a randomly mixed population, we assumed homogeneous and random mixing of individuals. For a given number of contacts, *n*, and vaccination rate, *p*, the probability of at least one individual being under or unvaccinated among the *n* contacts is given by the formula 1 – (1-*p*))^*n*^.

### Assessing the relative risk reduction of vaccination

To estimate the relative risk reduction of incremental vaccination, we first estimated the stochastic risk of an outbreak for a baseline scenario representing 90% vaccination rate. We then calculated the probability of an outbreak of 10 or more cases, representing a significant outbreak, at different population sizes, using this as the baseline risk of an outbreak. We then reran the simulation, increasing the percentage of the population that is fully vaccinated, allowing us to calculate a relative risk (RR) reduction for each marginal increase in the percentage vaccinated where RR reduction = 1 – probability of an outbreak with increased vaccination / probability of an outbreak with no increase in vaccination.

All analysis was conducted using R version 4.3.3. Data and code are available at https://github.com/wf-id/measles-risk.

## Results

Our analysis suggests that a reasonable estimate for the number of under or unvaccinated (at-risk) elementary-aged students in North Carolina is roughly 41,352 out of an estimated enrollment of 641,048 suggesting an aggregated 93.5% full vaccination rate. Only 47 of the 100 North Carolina counties have full vaccination rates of at least 95% (Figure 1A). We find that elementary-aged vaccination ranges from 87% in Cherokee County to 98% in Hyde County with the number of at-risk students ranging from 3 in Hyde County to over 5,670 in Mecklenburg County. The top 5 counties at risk represent over 40% of the total students at-risk (Figure 2B). Among the 22 non-rural counties, 23% have vaccination rates at or above 95%, compared to 54% of the 78 rural counties. Seventy-two percent of at-risk children (N = 29,795) reside in non-rural counties.

**Figure 2.**
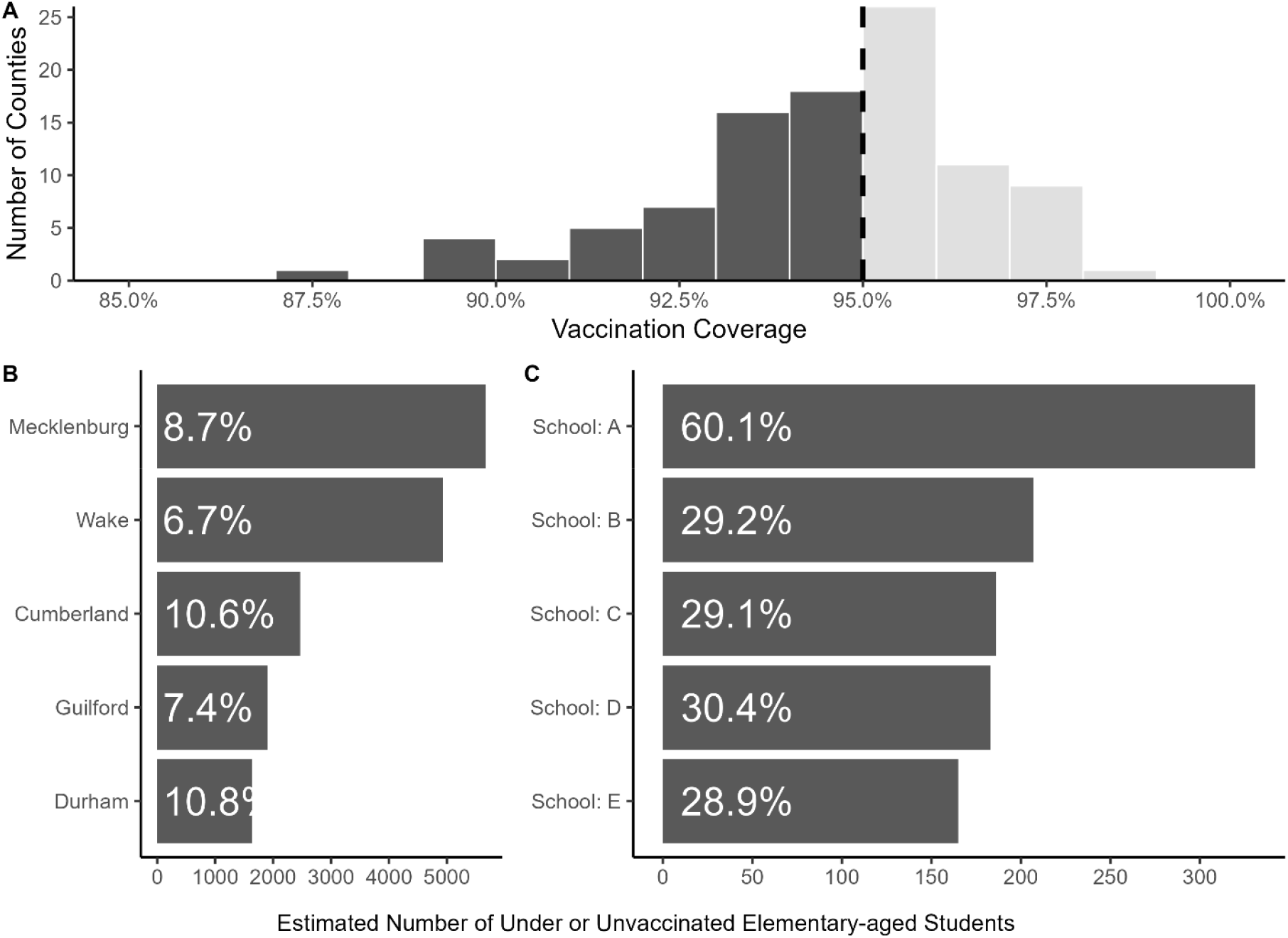
The distribution of North Carolina Counties by vaccination coverage (A). The North Carolina counties with the estimated largest number of at-risk students by number of students (B) and by school vaccination rates (C). For schools to appear, they must have more than 2 years of immunization data reported. The percentage represented the estimated vaccination rate for grades Kindergarten to 5^th^ grade.

When examining the vaccination rates by school, we find substantial heterogeneity within schools within the same counties. Among the 1,975 elementary schools, 1,474 (76%) had sufficient immunization reporting rates. Of the 1,474 schools, only 46% have vaccination rates at or above 95%. One school has a vaccination rate as low as 40%.

The schools with the top five estimated number of at-risk students and their vaccination rates are shown in Figure 2C. Full estimates for each school are available in the appendix.

Among the 1,474 elementary schools with sufficient data, we estimate that the average outbreak size among all schools is 7 cases (95% confidence interval, 0 to 22). However, 98 elementary schools have estimated average outbreaks of at least 20 students and 6 elementary schools with estimated outbreak sizes of at least 100 students (Figure 3A). Similarly, 142 elementary schools have over a 50% probability of at least 10 cases with 28 having a probability of over 80% (Figure 3B). The average elementary school (kindergarent-5^th^ grade) in North Carolina is estimated to have roughly 402 students and a full vaccination rate of 93%.

**Figure 3.**
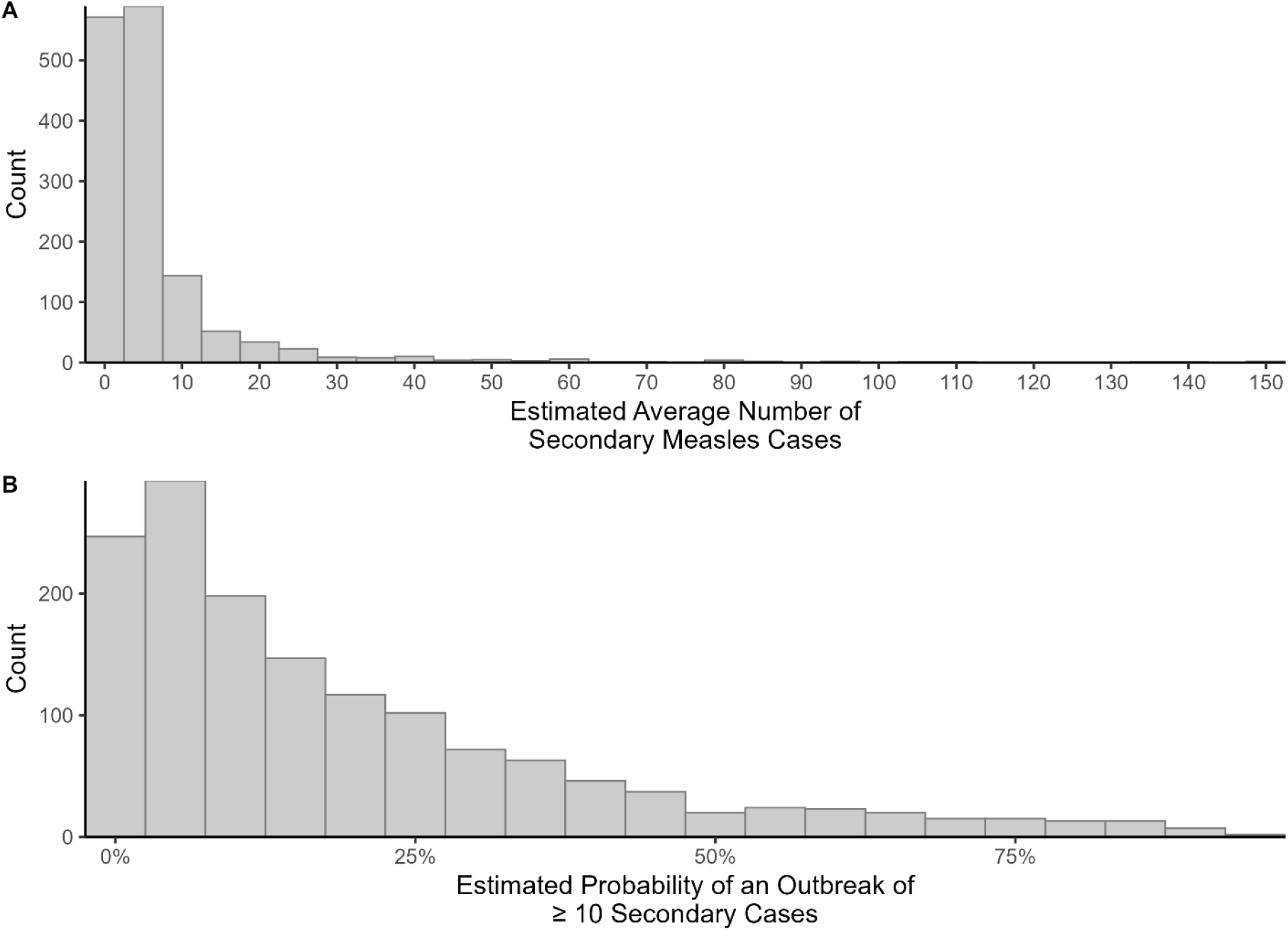
A histogram of the estimated average outbreak sizes for measles cases upon the introduction of a single infected student in North Carolina elementary schools (A). The distribution of estimated probabilities of outbreaks of at least 10 secondary measles cases among elementary school-age students in North Carolina, represented as the proportion of stochastic simulations which resulted in at least 10 cases (B).

When assessing the probability that an infected individual encounters an at-risk individual, we find that at current statewide average vaccination rates of 93.5% that if an individual had more than 10 encounters, there is a greater than 50% probability one encounter was with an at-risk individual (Figure 4A). Similarly, as vaccination rates decrease, then the probability that a contact is at risk increases. We found that vaccination not only decreases the probability that a contact is at risk, but also that a single percentage increase (i.e., from 90% to 91%) in vaccination coverage decreases the relative risk of an outbreak by roughly 20% (Figure 4B).

**Figure 4.**
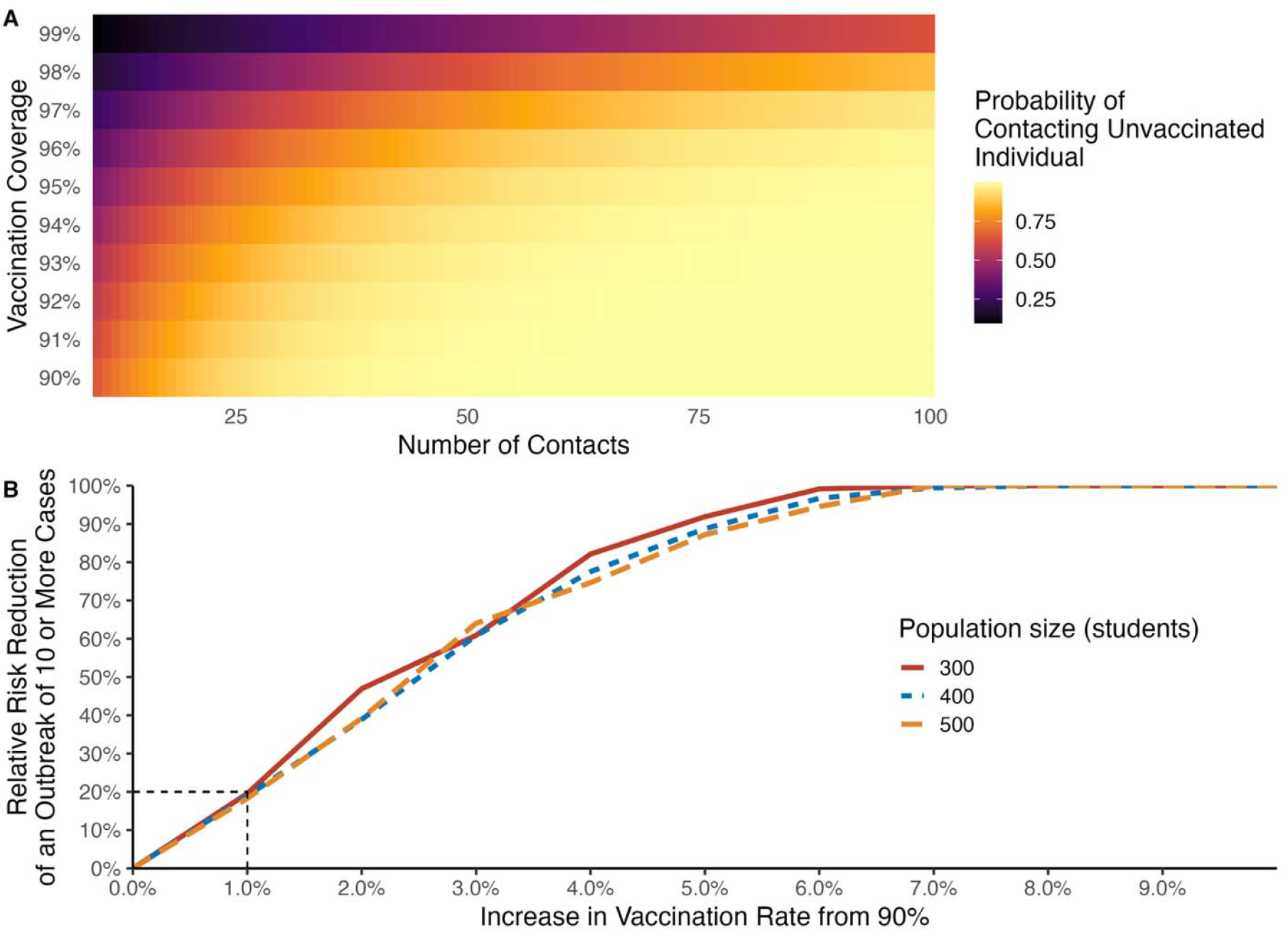
A heatmap showing the probability of an infected individual encountering at least one under vaccinated or unvaccinated individual assuming the number of contacts and that vaccinated individuals are distributed randomly (A). Lighter colors represent higher probabilities of contacts while darker colors represent lower probabilities. The relative risk reduction in the probability of an outbreak of ten or more students given the marginal increase in the student vaccination rate over 90% given three different population sizes (B). The dashed black lines represent the marginal risk reduction for increasing vaccination from 90% to 91%.

To better facilitate the use of these findings, we have generated a dashboard which displays these analysis and data for both public health officials and general consumption at https://measles2025abc.wakeforestid.com.

## Discussion

Our analysis, while representing a rough order of magnitude estimation of risk and outbreak probability, suggests that there is substantial risk for measles outbreaks across North Carolina. As vaccination rates across the United States continue to decline, there is a large risk that measles and other vaccine preventable illnesses increase in circulation and return to endemicity within the next 20 years[8]. The 2025 reported case is the second case of measles infection in North Carolina since 2024 after several years with no reported cases. These cases the ongoing national outbreaks of measles[1], and the pattern of reduced vaccination coverage together show that the risk of onward transmission is an imminent threat. The recent North Carolina case spent nearly five hours in a science center which has an annualized average of 1700 visitors per day. Our analysis would suggest that it is highly likely (effectively 100%) that the infected individual may have exposed at least one other under or unvaccinated individual assuming the vaccination coverage in Guilford county.

Substantial heterogeneity in measles vaccination rates exists across North Carolina counties and even within counties at different elementary schools. This heterogeneity in vaccination rates can further amplify the risk of onward transmission and increase the risk of larger outbreaks[22,23]. Furthermore, the existence of these pockets of unvaccinated and under vaccinated individuals, can help to serve as sources, seeding outbreaks and re-developing cycles of infection[24]. Unvaccinated individuals with measles are 3 to 4 times more infectious compared with vaccinated individuals with measles. In addition, unvaccinated primary measles cases are more likely to transmit the infection to unvaccinated people compared with transmission to vaccinated people.[25]

A 2007 measles outbreak investigation in Quebec, Canada revealed that despite a high average population vaccine coverage, there were 10 generations of transmission between unrelated unvaccinated networks of individuals[26]. Similarly, a single infected child in New York City in 2018 led to a total of 649 additional measles cases, underlying the consequences of even one infected individual in pockets of under and unvaccinated individuals[27].

Measles vaccination has led to a tremendous reduction in measles-related morbidity and mortality. Modeling studies estimate that 154 million deaths have been avoided between 1974 to 2024 as a consequence of vaccination[28]. Our calculations show that a single percentage increase in vaccination coverage (e.g., from 90% to 91%) in a 500-student school setting decreases the relative risk of an outbreak by roughly 20%. The vaccine is regarded as safe and well tolerated, generating high levels of protection against infection and symptomatic disease [14,29]. Measles infections, on the other hand, can induce negative effects on existing humoral immune memory[30] along with high rates of acute symptoms requiring medical care, in addition to potential long-term, potentially fatal sequala such as measles inclusion body encephalitis in immunocompromised hosts and subacute sclerosing panencephalitis[12].

Our study is not without limitations. We assumed that the 2019-2020 immunization records could be represented by the 2020-2021 metrics as well as the 2024-2025 by the 2023-2024 values. Deviations from this assumption will have an impact on the number of at-risk individuals we estimated. Vaccination rates in 2020 were identical at a state level to 2019 according to CDC data [31]; however, given the current trends in immunizations, our estimate for 2024-2025 may underestimate the number of elementary-aged students at risk. As only kindergarten data are available, we make the strong assumption that elementary-aged students do not receive these vaccines at a later time (e.g., students who were unvaccinated as kindergarteners 2 years ago are assumed to remain unvaccinated as second graders). Our models assume that the populations of the schools remain constant during an outbreak (i.e., no movement of students between the schools). Differential movement of infected and susceptible children could drive additional transmission [32]. Additionally, we assume that provisionally enrolled kindergarteners representing less than 2% of the total number of kindergartners in North Carolina, remain unvaccinated and within the at risk population group [9].

Our mathematical models assume that school populations are homogenously mixed. Given the dynamics of schools (e.g., shared resources like the media center, cafeterias, gyms) and the long airborne transmission potential of measles, we feel that this is a justified assumption. These compartmental models are standard for modeling measles outbreaks and other directly transmitted infections [10,33]. By executing the stochastic models for 1,000 iterations we captured the potential for “ stochastic die-off” in which a single case fails to spark a transmission chain. We hope that by capturing the range of outbreak scenarios that we provide reasonable estimated for the size and frequency of different outbreaks in different school’s sizes and vaccination rates.

Ultimately by making these data and analysis publicly available, we hope to provide stakeholders with information to assess both public health response and understand personal risk. These results may inform local public health department to target MMR vaccination strategies at high-risk countries in our state. Additionally, with publicly available data we hope that trusted community partners, providers, and pediatricians can have additional information to discuss with the communities that they serve. These rough, order-of-magnitude estimates can provide public health officials the ability to identify areas of low vaccination and higher risk of outbreak such that they can adequately plan for outbreak response and prioritize school and communities at risk for vaccination campaigns.

## Data Availability

Data and code are available at https://github.com/wf-id/measles-risk.

https://measles2025abc.wakeforestid.com

## Acknowledgements

Computations were performed using the Wake Forest University (WFU) High Performance Computing Facility, a centrally managed computational resource available to WFU researchers including faculty, staff, students, and collaborators.

